# Childhood intelligence and risk of depression in later-life: A longitudinal data-linkage study

**DOI:** 10.1101/2023.08.08.23293657

**Authors:** Emily L Ball, Drew M Altschul, Simon R Cox, Ian J Deary, Andrew M McIntosh, Matthew H Iveson

**Affiliations:** Division of Psychiatry, Centre for Clinical Brain Sciences, University of Edinburgh, Edinburgh, UK; Department of Psychology, University of Edinburgh, Edinburgh, UK; Lothian Birth Cohorts, Department of Psychology, University of Edinburgh, Edinburgh, UK

**Keywords:** Depression, Intelligence, Environmental risk factors, Cognitive epidemiology

## Abstract

**Background:** Lower childhood intelligence test scores are reported in some studies to be associated with higher risk of depression in adulthood. The reasons for the association are unclear. This longitudinal data-linkage study explored the relationship between childhood intelligence (at age ∼11) and risk of depression in later-life (up to age ∼85), and whether environmental factors in childhood and adulthood accounted for some of this association.

**Methods:** Intelligence test scores collected in the Scottish Mental Survey 1947 were linked to electronic health records (hospital admissions and prescribing data) between 1980-2020 (n=53,037), to identify diagnoses of depression. Mixed-effect Cox regression models were used to explore the relationship between childhood intelligence test scores and risk of depression in later-life. Analyses were also adjusted for environmental factors experienced in childhood (number of siblings) and adulthood (Carstairs index, urban/rural).

**Results:** Twenty-seven percent of participants were diagnosed with depression during follow-up (n=14,063/53,037). Greater childhood intelligence test scores were associated with a reduced risk of depression in an unadjusted analysis (HR=0.95, 95% CI=0.93 to 0.97, P<0.001), and after adjustment for environmental factors experienced in childhood and adulthood (HR=0.95, 95% CI=0.91 to 1.00, P=0.032).

**Conclusions:** This study provides additional evidence of an association between higher childhood intelligence and reduced risk of later depression. Of the environmental factors included in this study, childhood and adulthood environmental factors did not seem to be substantial confounders.

**Key messages:**

- Identifying modifiable environmental risk factors, may help to identify interventions for the primary prevention of depression.
- Greater childhood intelligence test scores (at age 11) were associated with a reduced risk of depression in later-life (up to age 85) following unadjusted analysis.
- The association between childhood intelligence and risk of depression in later-life remained even after adjusting for childhood environmental factors (number of siblings) and adulthood environmental factors (Carstairs and urban/rural).
- How depression is defined in epidemiological research (e.g., diagnostic codes in hospital admissions or being prescribed antidepressants) could influence the association between childhood intelligence and depression in later-life.

## Introduction

Depression is estimated to affect over 280 million people worldwide.^1^ Uncovering ways to prevent depression is the main research priority for people who have experienced depression (James Lind Alliance Priority Setting Partnerships).^2^ There are many risk factors for depression including sex, genetic, and environmental factors.^3^ Many environmental factors originate in early life. Identifying modifiable environmental risk factors, particularly in childhood, may help to identify interventions for the primary prevention of depression.

Intelligence is also associated with risk of depression in many, but not all, studies. A meta-analysis of 34 study samples (categorical and continuous outcomes of depression) found that higher intelligence decreased the risk of subsequent depression (r=-0.09, 95%CI=-0.121 to -0.054, P<0.001).^4^ However, this association was confounded by the presence of depressive symptoms at the time of cognitive assessment.^4^ Of the 23 studies in the review that assessed cognition prior to age 18 years, none measured the association between childhood intelligence and depression after age 55 years. A subsequent study found that higher intelligence (assessed between age 15-23 years) was associated with an increased risk of depression prior to age 50, but a lower risk of depression at the age of 50 years.^5^ These findings highlight that when trying to understand the association between intelligence and depression, and the potential causes of the association, studies should follow-up participants throughout the life course.

Several studies have reported that lower childhood intelligence predicts premature mortality and comorbidities (e.g., cardiovascular disease).^6–8^ Having a measure of intelligence in childhood is highly valuable as it allows researchers to understand the association between intelligence and subsequent health outcomes without the potentially confounding effects of contemporaneous depressive symptoms and subsequent level of education (lower level of education is also reported to increase risk of later-life depression^9^).

On the 14^th^ June 1947, nearly all children aged ∼11 years in Scotland took part in the Scottish Mental Survey 1947 (SMS1947). This survey assessed intelligence using the Moray House Test No. 12.^10^ A subsequent study, examined the relationship between childhood intelligence and symptoms of depression in later-life (assessed at five intervals between the years of 2004 (n=1091) and 2019 (n=431)) in a subset of SMS1947.^11^ This study did not find an association between childhood intelligence and risk of depression, however, they only measured depressive symptoms experienced a week prior to the follow-up intervals, meaning cases of depression could have been missed. In our current study, we included a much larger sample of participants who had taken part in SMS1947 (n=53,037), we also improved the coverage of depression by identifying cases of depression reported in electronic health records, thus reducing the risk of attrition.

As described above, there is a well-established association between early-life intelligence and later occurrence of depressive illness, however, we do not understand the causes of this association. Given findings that higher household income, higher education, and urbanisation increase risk of depression,^12,13^ and these factors relate to both intelligence and depression, it is important to account for these potentially important environmental factors across the life course. As presented in Figure 1, environmental factors experienced in childhood (e.g., parental income, family structure, geographical location, emotional/social support) and adulthood (e.g., income, social class, work-stress) may mediate or moderate the association between childhood intelligence and depression in later-life. For example, having fewer siblings in childhood may enable a child to receive more financial support from their parents, which could lead to better educational and recreational opportunities, and a higher paid job in adulthood. These factors may reduce a person’s risk of depression.

**Figure 1:**
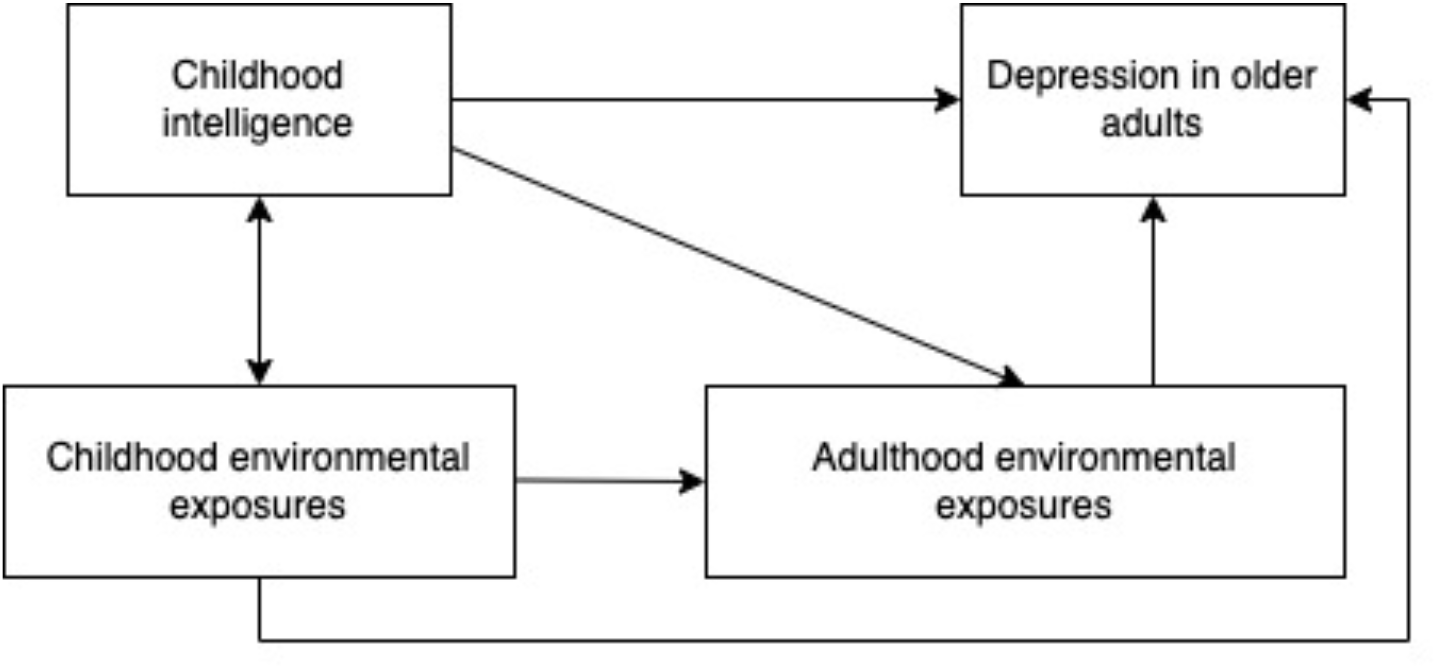
Schematic diagram presenting possible interactions between childhood intelligence, environmental factors across the life course and depression in older adults.

Intelligence may also influence the environmental factors that a person is exposed to (i.e., those with higher intelligence may be exposed to better socioeconomic conditions, that may in turn minimise the risk of depression). Thus, the present well-powered population-level study (using a year-of-birth cohort in all of Scotland; n>53,000) with a measure of cognitive differences in early life (prior to secondary-level educational differences and depression onset), environmental factors in childhood and adulthood, health record data linkage, and an exceptionally long follow up period into older age (up to age 85 years) might offer important insights into the relationship between childhood intelligence and risk of depression in later-life.

## Methods

This study is reported according to the RECORD checklist.^14^

### Study population

The study population was formed of participants who took part in the Scottish Mental Survey in 1947 (n=70,805).^10^ The Scottish Mental Survey 1947 (SMS1947) assessed intelligence of almost all children who were born in Scotland in 1936 and attended a Scottish school on 4th June 1947.^8,15^ Intelligence was assessed using the Moray House Test No. 12, and information including sex, number of siblings, and position in the family (e.g. first born child) was also collected.^15^

Using the participants’ Community Health Index (CHI) numbers (a 10-digit identifier recorded in electronic health records), data collected in SMS1947 were linked to electronic health records from later in life. Participants were excluded from the study population when their electronic health records could not be traced (e.g., they may no longer live in Scotland or died prior to the start of follow-up). Follow-up for diagnoses of depression began in 1980 to 2020.

### Data linkage to electronic health records

The SMS1947 cohort were linked to the following electronic health records: General/Acute Inpatient and Day Case (SMR01 – data available from 1980 to 2020), Mental Health Inpatient and Day Case (SMR04 – data available from 1980 to 2020), the Prescribing Information System (PIS – data available from 2006 to 2020 – largely available from 2009), and National Records of Scotland Deaths (1970 to 2020). Sex was taken from Community Health Index records generated from first contact with an NHS service (usually birth). Data were linked by the eDRIS team (Public Health Scotland) and pseudonymised datasets were provided to researchers for analysis within a Trusted Research Environment.

### Outcome

Through linkage of SMS1947 data to electronic health records, we identified whether participants had received a diagnosis of depression after age ∼44 years (when electronic health records became available). We searched for diagnoses of depression in ‘main’ or ‘other’ conditions in hospital admission records (Scottish Morbidity Records; ICD codes presented in Supplementary Table S1) and for antidepressants that had been prescribed for a minimum period of six consecutive weeks (Prescribing Information System; list of antidepressants presented in Supplementary Table S2). Individuals without a depression hospital admission or antidepressant prescription are defined as ‘no depression’.

### Environmental factors

Childhood cognitive test scores and environmental factors analysed in this study were collected at age 11 (childhood intelligence, position in the family, number of siblings) (Table 1). Adulthood environmental factors relating to socioeconomic status (SES) were collected at the time of a hospital admission (Table 1). Adulthood environmental factors were not available at the time that antidepressants were prescribed. Environmental factors (such as the location a person lives) are likely to remain stable within a short period of time, for example, data collected in the 2011 census highlighted that the probability of relocation within Scotland is highest in young adults and significantly declines with age.^16^ If a person was prescribed antidepressants within six months of a hospital admission (including non-depression admissions), we included the adulthood environmental factors from the most recent hospital admission in the analyses of antidepressant data.

**Table 1:** Description of risk factors.

|  | Risk factor | Definition | When collected | How it was treated in statistical models |
| --- | --- | --- | --- | --- |
|  | **Sex** | Male<br>Female | Community Health Index records | Categorical variable |
|  | **Childhood Intelligence** | Moray House Test No. 12 Score<sup>10</sup> | At age 11 | Continuous variable – corrected for participant age (to account for small differences in age at test). The variable was also IQ-scaled (M=100, SD=15) and standardised (M=0, SD=1), so that estimates are based on a 1SD (15-IQ-point) change |
| **Childhood environment** | **Position in the family** | Position within number of siblings (e.g., first born, second born) | At age 11 | Continuous variable – higher numbers mean later birth order |
|  | **Number of siblings** | Number of siblings | At age 11 | Continuous variable – Yeo Johnson transformed due to non-linearity |
| **Adulthood environment** | **Carstairs deprivation index decile**<br>(Classified according to 2001 criteria) | Carstairs index is comprised of: car ownership, male unemployment, overcrowding and low social class.<sup>17</sup> (Carstairs index is a decile because areas in Scotland are ranked against other areas in Scotland). | At hospital admission | Continuous variable – higher numbers mean more deprived |
|  | **Urban/rural classification**<br>(Classified according to 2003/2004 criteria) | Urban ≥ 3000 people<br>Rural < 3000 people<sup>18</sup> (categorised using area-based measures e.g., population size and accessibility) | At hospital admission | Binaries according to the Scot Gov 2-fold class system<sup>18</sup> |
|  | **Scottish Index of Multiple Deprivation rank**<br>(Classified according to 2004 criteria) | Formed of 8 ranks, ranking areas of deprivation in Scotland<sup>19</sup> | At hospital admission | Ordered categorical variable |

### Statistical analysis

Analyses were conducted using R (Version 4.2.0).^20^ We presented descriptive statistics categorised by sex. Kruskal-Wallis tests were conducted for numeric variables and chi-squared tests were conducted for categorical variables. Prior to including variables in a regression model, we checked multicollinearity between childhood and adulthood environmental risk factors (position in the family, number of siblings, Carstairs, urban/rural, SIMD), tested the proportional hazards assumption, checked for influential observations, and checked for non-linearity. Since ‘number of siblings’ was non-linear we used the Yeo-Johnson transformation of the variable in the statistical models. Participants were censored at the time of depression diagnosis, at death, or at the end of follow-up in 2020.

A proportion of participants had multiple diagnoses of depression (repeated outcome measure). For each participant, we included all hospital admissions containing a diagnosis of depression and/or the earliest time they had been prescribed antidepressants (for at least six consecutive weeks) in our main analyses. To account for within and between participant variability, and thus random error, we performed mixed-effects models. We performed unadjusted mixed-effects Cox models (coxme package)^21^ to assess the association between childhood and adulthood environmental factors and depression in later-life. We then further explored the association between childhood intelligence and later-life depression by adjusting for sex and: (Model 1) childhood environmental factors (position in the family, number of siblings), (Model 2) adulthood environmental factors (Carstairs, urban/rural), (Model 3) all environmental factors (position in the family, number of siblings, Carstairs, urban/rural). Since female sex is a known risk factor for depression, we also stratified the analyses by sex.^3,22^

Depression identified in hospital admissions records, particularly when depression was the ‘main’ reason for admission, are likely to reflect more severe cases of depression. We therefore performed two sensitivity analyses which involved: (1) identifying cases of depression using only prescribed drugs data, (2) identifying cases of depression using only hospital admissions data.

### Ethics

The study was reviewed by an NHS Research Ethics Committee who waived the need for full ethical review of this work (18/SC/0505).

## Results

### Participant characteristics

70,805 11-year old participants undertook the Moray House Test No. 12 in 1947. 17,768/70,805 (25%) participants were excluded from our study because it was not possible to trace their electronic health records, reasons for being unable to trace records include no longer living in Scotland or dying prior to the start of follow-up. Table 2 presents descriptive statistics for the study sample (n=53,037). Adult environmental risk factors (Carstairs and urban/rural) are only available for participants who had a hospital admission (with/without a diagnosis of depression); hence, these variables are missing for those with no hospital admission.

**Table 2:** Descriptive statistics relating to the study sample.

| Risk factors | Females<br>(n=26,803) | Males<br>(n=26,234) | Total<br>(n=53,037) | P value |
| --- | --- | --- | --- | --- |
| **Moray House Test score at age ~11 years-old**<br>**(Variable not age corrected or standardised, max possible score = 76)** |  |  |  | **<0.001** |
| Mean (SD) | 37.39 (14.98) | 35.37 (16.36) | 36.39 (15.71) |  |
| n Missing | 1773 | 1753 | 3526 |  |
| **Position among children in family at age 11 years-old** |  |  |  | **0.057** |
| Mean (SD) | 2.55 (1.85) | 2.51 (1.80) | 2.53 (1.82) |  |
| n Missing | 269 | 226 | 495 |  |
| **Number of children in family at age 11 years-old** |  |  |  | **0.079** |
| Mean (SD) | 3.83 (2.25) | 3.78 (2.20) | 3.80 (2.22) |  |
| n Missing | 272 | 227 | 499 |  |
| **Carstairs decile at admission (2001 scheme)±** |  |  |  | **<0.001** |
| Mean (SD) | 5.95 (2.86) | 6.16 (2.87) | 6.06 (2.86) |  |
| n Missing | 20,557 | 19,720 | 40,277 |  |
| **Urban-rural classification at admission (2003/2004 scheme)\*±** |  |  |  | **0.229** |
| Accessible | 5259 (84.9%) | 5428 (84.1%) | 10687 (84.5%) |  |
| Remote | 937 (15.1%) | 1026 (15.9%) | 1963 (15.5%) |  |
| n Missing | 20,607 | 19,780 | 40,387 |  |
| **SIMD classification at admission (2004 scheme)±** |  |  |  | **<0.001** |
| Mean (SD) | 6.09 (2.86) | 6.37 (2.80) | 6.24 (2.84) |  |
| 1 | 478 (7.7%) | 376 (5.8%) | 854 (6.7%) |  |
| 2 | 472 (7.6%) | 427 (6.6%) | 899 (7.1%) |  |
| 3 | 519 (8.4%) | 478 (7.4%) | 997 (7.9%) |  |
| 4 | 479 (7.7%) | 516 (8.0%) | 995 (7.8%) |  |
| 5 | 572 (9.2%) | 609 (9.4%) | 1181 (9.3%) |  |
| 6 | 627 (10.1%) | 671 (10.4%) | 1298 (10.2%) |  |
| 7 | 693 (11.2%) | 707 (10.9%) | 1400 (11.0%) |  |
| 8 | 761 (12.2%) | 774 (11.9%) | 1535 (12.1%) |  |
| 9 | 739 (11.9%) | 850 (13.1%) | 1589 (12.5%) |  |
| 10 | 875 (14.1%) | 1069 (16.5%) | 1944 (15.3%) |  |
| n Missing | 20,588 | 19,757 | 40,345 |  |
| **Age at first diagnosis of depression (hospital admission/antidepressants)±** |  |  |  | **<0.001** |
| Mean (SD) | 74.42 (6.34) | 74.70 (6.44) | 74.52 (6.38) |  |
| Total n | 8826 | 5237 | 14,063 |  |
| **Age earliest antidepressant prescription that lasted >6 weeks** |  |  |  | **<0.001** |
| Mean (SD) | 75.76 (3.20) | 76.22 (3.23) | 75.93 (3.22) |  |
| Total n | 8348 | 4834 | 13,182 |  |
| **Age at earliest hospital admission for depression** |  |  |  | **0.825** |
| Mean (SD) | 65.59 (11.26) | 65.82 (10.90) | 65.68 (11.12) |  |
| Total n | 1268 | 808 | 2076 |  |
| **Age at censor if they do not have depression (death or end of study)** |  |  |  | **<0.001** |
| Mean (SD) | 77.73 (10.17) | 75.32 (10.85) | 76.44 (10.61) |  |
| Total n | 17977 | 20997 | 38974 |  |
| **Depression** |  |  |  | **<0.001** |
| No depression | 17977 (67.1%) | 20997 (80.0%) | 38,974 (73.5%) |  |
| Depression | 8826 (32.9%) | 5237 (20.0%) | 14,063 (26.5%) |  |
| **Alive/died at end of follow-up** |  |  |  | **<0.001** |
| Alive | 14,419 (53.8%) | 10,521 (40.1%) | 24,940 (47%) |  |
| Deceased | 12,384 (46.2%) | 15,713 (59.9%) | 28,097 (53.0%) |  |

\*8-class scale which has been binarized
±Based on wide-format data table (1 row per participant)
n: number; SD: standard deviation;
Kruskal-Wallis tests are conducted for numeric variables and chi-squared tests are conducted for categorical variables.
p-value tests sex differences

### Depression cases

Twenty-seven percent (n=14,063/53,037) of participants had a diagnosis of depression reported in hospital admissions data and/or had been prescribed antidepressants for a consecutive period of six weeks or more. Most depression diagnoses (n=11,987/14,063; 85%) were identified using antidepressant prescribing data only (Prescribing Information System) (Figure 2). Six percent (n=881/14,063) of depression cases were identified in hospital admissions data only (Scottish Morbidity Records), and 8% (n=1195/14,063) of participants had been prescribed antidepressants and had a depression diagnosis reported in hospital admission records (Figure 2). Most depression diagnoses reported in hospital admissions were reported as ‘other’ diagnoses, rather than the main reasons for admission (Supplementary Figure S3). This means that a majority of people were admitted to hospital for another physical or mental condition but also had a diagnosis of depression listed as a secondary condition.

**Figure 2:**
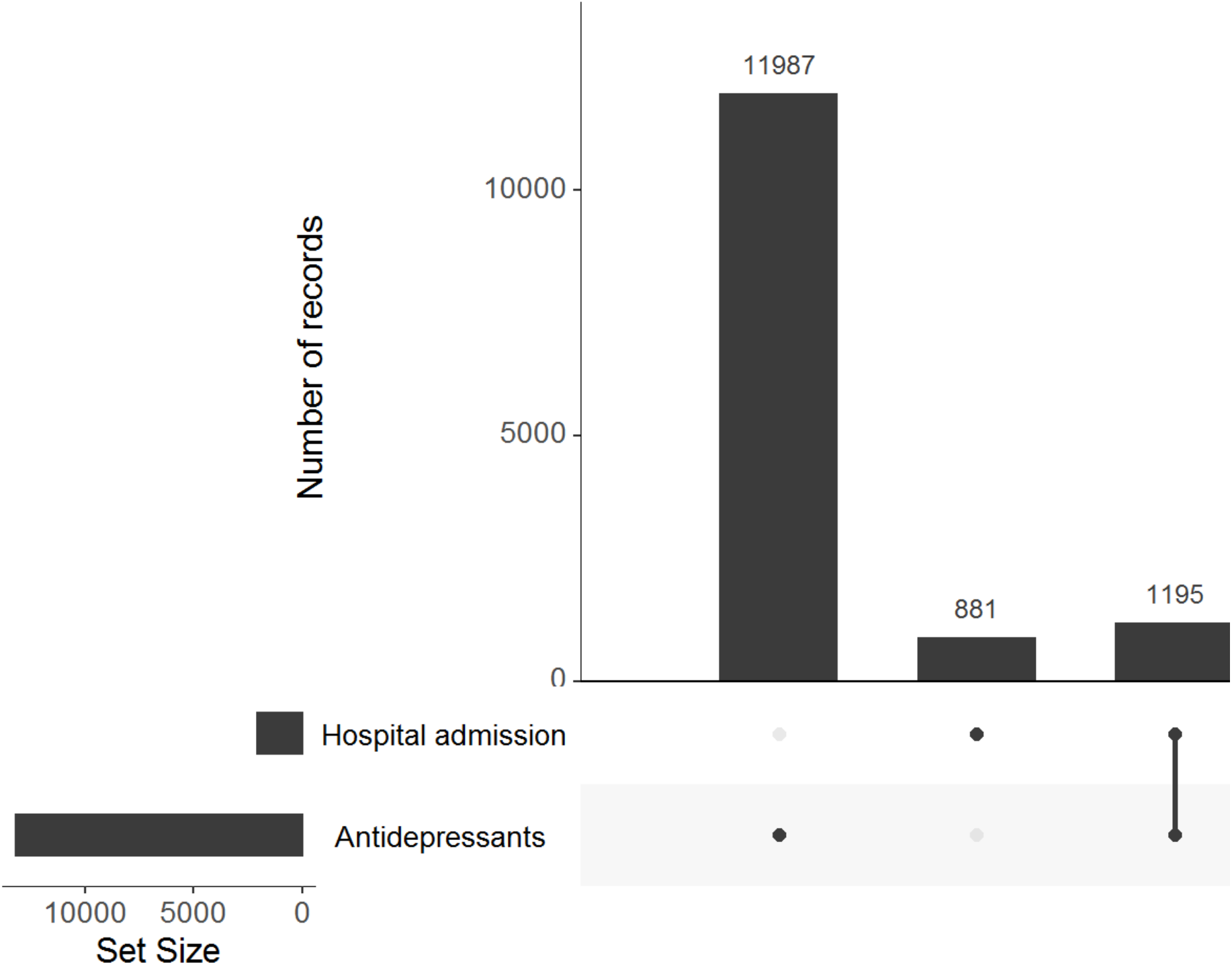
How many people were identified as having depression in hospital admissions and/or prescribed drugs data? The plot shows the number of participants who were either prescribed antidepressants (for a consecutive period of at least six weeks) and/or had a diagnosis of depression reported in hospital admissions records. The dots show the combinations of the sources (hospital admission/antidepressants).

### Univariable analysis

Following unadjusted mixed-effects Cox regression, being male (HR=0.68, 95% CI=0.66 to 0.71, P<0.001) and having a 1SD advantage in childhood intelligence (HR=0.95, 95% CI=0.93 to 0.97, P<0.001) were associated with a reduced risk of depression in later-life (Table 3 & Supplementary Table S4). Having a larger family size at age 11 (HR=1.03, 95% CI = 1.01 to 1.05, P=0.012), and having a higher Carstairs score (HR=1.01, 95% CI=1.00 to 1.03, P=0.048) were associated with an increased risk of depression (Table 3).

**Table 3:** Risk factors associated with depression after unadjusted and adjusted analyses (mixed effect models)

|  | Unadjusted risk factors<br>(Depression n/Total n) | Model 1:<br>Adjusted for childhood<br>risk factors | Model 2:<br>Adjusted for adulthood<br>risk factors | Model 3:<br>Adjusted for all<br>risk factors |
| --- | --- | --- | --- | --- |
|  | HR<br>(95% CI) | HR<br>(95% CI) | HR<br>(95% CI) | HR<br>(95% CI) |
| **Depression**<br>**n/Total n (%)±** | N/A | 16595/96102<br>(17%) | 5637/57341<br>(10%) | 5537/56771<br>(10%) |
| **Male** | 0.68<br>(0.66 to 0.71)<br>P<0.001<br>(n=18043/104762) | 0.68<br>(0.65 to 0.71)<br>P<0.001 | 0.84<br>(0.77 to 0.92)<br>P<0.001 | 0.84<br>(0.77 to 0.92)<br>P<0.001 |
| **Moray House Test** | 0.95<br>(0.93 to 0.97)<br>P<0.001<br>(n=16781/96960) | 0.94<br>(0.92 to 0.96)<br>P<0.001 | 0.95<br>(0.91 to 1.00)<br>P=0.035 | 0.95<br>(0.91 to 1.00)<br>P=0.032 |
| **Position in family** | 1.01<br>(1.00 to 1.02)<br>P=0.142<br>(n=17813/103605) | N/A | N/A | N/A |
| **Size of family** | 1.03<br>(1.01 to 1.05)<br>P=0.012<br>(n=17810/103591) | 1.00<br>(0.98 to 1.03)<br>P=0.709 | N/A | 1.00<br>(0.96 to 1.05)<br>P=1.000 |
| **Carstairs** | 1.01<br>(1.00 to 1.03)<br>P=0.048<br>(6165/63163) | N/A | 1.01<br>(1.00 to 1.03)<br>P=0.200 | 1.01<br>(0.99 to 1.03)<br>P=0.252 |
| **Remote location\*** | 1.05<br>(0.94 to 1.18)<br>P=0.350<br>(n=6117/62376) | N/A | 1.09<br>(0.97 to 1.22)<br>P=0.165 | 1.09<br>(0.97 to 1.22)<br>P=0.160 |
| **SIMD – Linear** | 1.12<br>(0.99 to 1.28)<br>P=0.077<br>(n=6127/62649) | N/A | N/A | N/A |

CI: confidence interval; HR: hazards ratio; SIMD: Scottish Index of Multiple Deprivation
± Mixed effects model is based on number of observations
\*(Accessible Rural Areas, Remote Rural Areas, Very Remote Rural Areas
The number of records included in each statistical model varies because records with missing environmental factors are excluded from the analyses.

We then assessed the correlations between cognitive test score and potential covariates prior to including in the adjusted models. All risk factors were correlated (P<0.001). As shown in Figure 3, the most strongly correlated covariates are number of siblings and position in the family, and, Carstairs and SIMD. We therefore excluded position in the family and SIMD from subsequent multivariable analyses.

**Figure 3:**
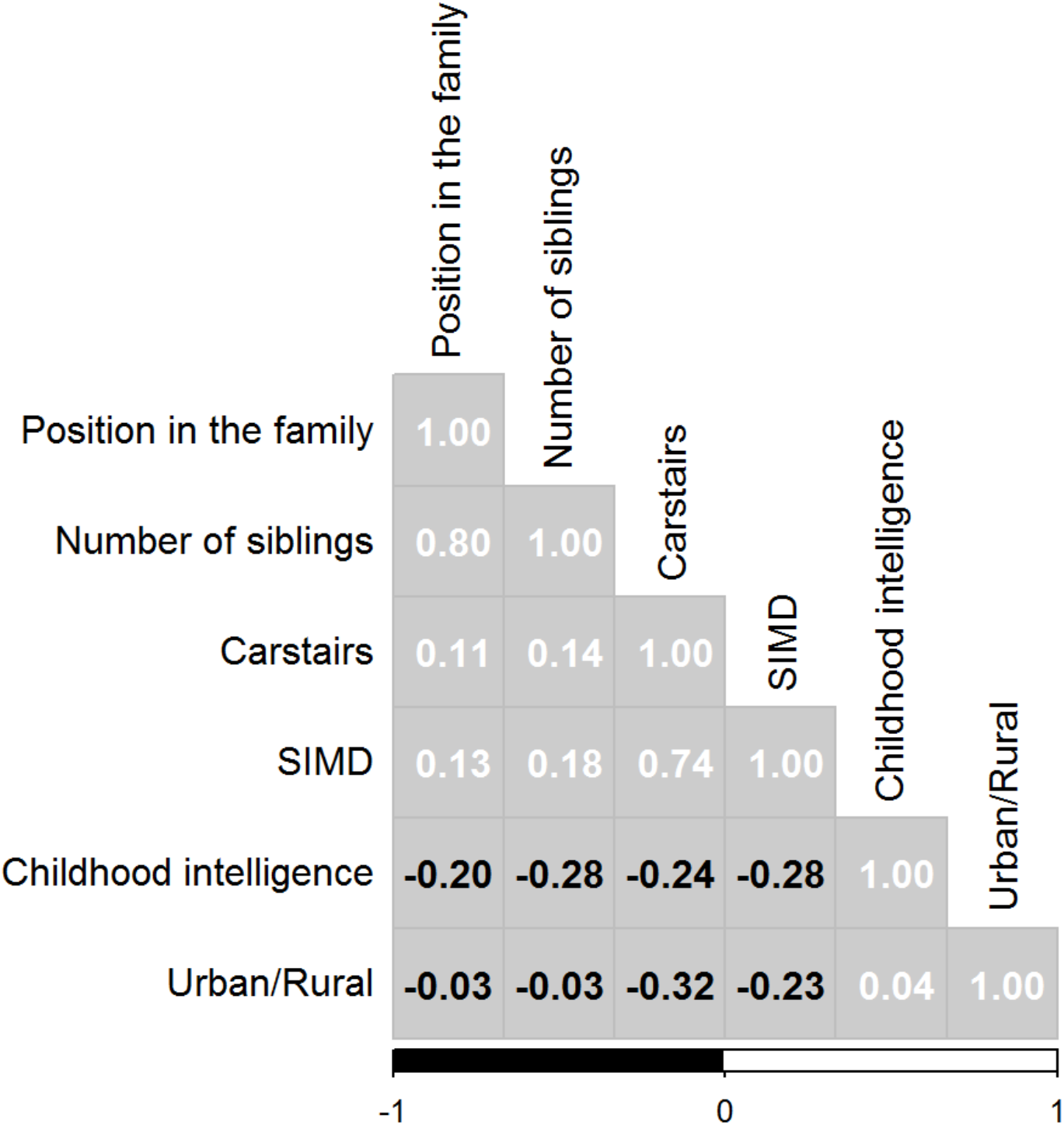
Covariate correlation matrix. Black numbers indicate a negative correlation SIMD: Social Index of Multiple Deprivation

### Multivariable analysis

After adjusting for sex and environmental factors in childhood (number of siblings), being male (HR=0.68, 95% CI=0.65 to 0.71, P<0.001) and a 1SD advantage in childhood intelligence (HR=0.94, 95% CI=0.92 to 0.96, P<0.001) was associated with reduced risk of depression (Table 3, Model 1).

When accounting for sex and environmental risk factors experienced in adulthood (Carstairs decile, urban/rural classification), a 1SD advantage in childhood intelligence was significantly associated with risk of depression (HR=0.95, 95% CI=0.91 to 1.00, P=0.035) (Table 3, Model 2). As in the unadjusted model, being male (HR=0.84, 95% CI=0.77 to 0.92, P<0.001) was associated with a reduced risk of depression in later-life (Table 3, Model 2).

Model 3 accounted for sex, and childhood and adulthood environmental risk factors. Childhood intelligence remained significantly associated with risk of depression (HR=0.95, 95% CI=0.91 to 1.00, P=0.032) (Table 3, Model 3). Male sex was associated with a reduced risk of depression (HR=0.84, 95% CI=0.77 to 0.92, P<0.001). Carstairs score or living in a rural location was not associated with risk of depression.

#### Sex stratified analyses

Stratifying by sex did not influence findings from unadjusted or adjusted analyses (Supplementary Table S5). The strength of association between cognitive score at age 11 and depression in later-life was similar in females (HR=0.94, 95% CI=0.92 to 0.97, P<0.001) and males (HR=0.93, 95% CI=0.90 to 0.96, P<0.001).This was also true for the other risk factors tested, although significance levels differed between females and males for Carstairs decile and SIMD (Supplementary Table S6).

#### Sensitivity analyses

When identifying cases of depression using only prescribed drugs data, we did not find any association between cognitive test score at age 11 and depression in later-life (Supplementary Table S7). However, when identifying cases of depression using only hospital admissions data, we found an association between childhood cognitive test score and depression following unadjusted analysis (HR=0.86, 95% CI=0.82 to 0.90, P<0.001), and after adjusting for environmental factors experienced in childhood and adulthood (HR=0.94, 95% CI=0.89 to 0.99, P=0.026) (Supplementary Table S8).

## Discussion

This longitudinal data-linkage study explored the relationship between childhood intelligence and risk of depression in later-life. Greater childhood intelligence was associated with a reduced risk of depression following unadjusted analyses. In addition to the conclusions in Scult et al., 2017, we also provide evidence of an intelligence-depression association which is present at age 11 and not likely to be confounded by concurrent depression symptoms nor by later-life factors such as education. The association between childhood intelligence and depression remained after adjusting for childhood environmental factors (number of siblings) and environmental factors experienced in adulthood (urban/rural classification, Carstairs deprivation). In agreement with the existing literature, male sex was consistently associated with a reduced risk of depression across later-life.^3^

Our findings differ from those of Wraw et al., 2016, who reported that higher early-life intelligence (between age 15-23) increased the risk of lifetime depression (up to age 50), and this association was strengthened after adjusting for adult SES (OR = 1.32, 95% CI = 1.16 to 1.51).^5^ Wraw et al., 2016 were able to look at the independent effects of three adult SES variables: adult educational attainment, income, occupational status, finding that income was the main facilitator strengthening this association.^5^ However, their population consisted of people who lived in the United States, and due to cost of health insurance, people with a higher income may be more likely to be able to seek a diagnosis of depression.

When conducting epidemiological research, it is important to consider how defining depression could influence the association between cognitive ability and depression. In addition to measuring risk of lifetime depression (up to age 50), Wraw et al., 2016, also measured depression using the 7-item Centre for Epidemiological Studies Depression Scale (CES-D).^5^ Following this analysis, they found that higher early-life intelligence was associated with less CES-depression.^5^ In our study, we identified cases of depression using hospital admissions data and prescribed drugs data. Our sensitivity analyses found there was no association between childhood intelligence and risk of depression when depression was identified using only prescribed drugs data. Conversely, when depression was identified in hospital admissions data, there was a stronger association between higher childhood intelligence test score and reduced risk of depression in later life. The stronger association between childhood intelligence and depression diagnosed in hospital admissions data could signal an interplay between childhood intelligence, physical health, and risk of depression. Due to the availability of data, depression reported in hospital admissions was also identified over a wider age range then depression identified in the Prescribing Information System.

Studies that have measured the association between intelligence and depression use heterogeneous methods. Previous studies have used a variety of tools to assess intelligence (e.g., general intelligence, specific cognitive domains), implemented various methods to identify cases of depression (e.g., self-report, interview), and report various effect sizes (e.g., odds ratio, hazards ratio, correlation coefficients).^4^ One large-scale study of 666,804 Danish men reported that cognitive ability (at mean age of 19 years) was associated with an increased risk of a single depressive episode when followed up for over 50 years, after stratifying by age and adjusting for educational level.^23^ To enable comparison with our findings, we calculated the inverse hazard ratio for this association (HRi = 0.84, 95% CIi = 0.82 to 0.86). The effect size reported by Christensen et al., 2018 is similar to the size of effect we found when only identifying cases of depression in hospital admissions data.^23^

Of the studies which were included in the systematic review by Scult *et al*., 2017, the largest sample size that assessed intelligence before the age of 18 years was 14,322 participants (mean age:15.98) and the maximum length of follow-up was 45 years.^4^ The longer length of follow-up in our well-powered study enabled us to look at the impact of childhood intelligence on risk of depression in later-life, whilst also exploring how childhood and adulthood environmental factors influence this relationship.

There are challenges with calculating the prevalence of depression in urban and rural areas. Living in a rural area may make it more difficult to access primary and secondary care for depression. This may lead to a higher prevalence of undiagnosed depression in rural areas which have not been detected in this study. We may have also missed cases of depression when affected people sought alternative treatments for depression (e.g., talking therapy) which are likely to be more easily accessible to those who live in urban areas. In agreement with the findings of our study, a systematic review published in 2019 pooled 18 studies (n=31,598) and reported that prevalence of depression did not differ between people who lived in rural and urban areas.^12^ There was also no significant association when the 18 studies were pooled in a meta-analysis (OR=1.18, 95% CI=0.84 to 1.65; an OR greater than 1 indicates an association between urban location and depression), however, the studies were highly heterogeneous (*I*^2^=93.4%, X^2^=P<0.001).^12^ Our study assessed the association between urban/rural living and risk of depression in a larger sample size but was limited to people living in Scotland. Understanding the interaction between living in an urban or rural area and risk of depression is complex and likely to be influenced by country and culture.

### Strengths and limitations

Key strengths of this study are the large sample size and long length of follow-up (over 85 years), combined with access to early-life intelligence, before potential confounding by downstream factors (e.g., education, occupation). The original SMS1947 sample captured almost an entire population of children, regardless of sex or background, and therefore avoids participation bias. Linking to electronic health records is also less resource intensive than following up participants in-person to diagnose depression and limits participant attrition. Nevertheless, people who have not been admitted to hospital, those who have sought alternative treatments (e.g., counselling), or have not sought clinical help will not be captured by our analyses. It has been widely reported that many people living with depression do not seek help; which is pronounced in males.^24^ Those admitted to hospital with a ‘main’ diagnosis of depression likely represent those living with the most severe symptoms, missing those with milder and sub-clinical symptoms.

Data-linkage methods provide a quicker, cheaper, less resource intensive method of identifying people with depression. Due to being unable to access primary care data, where most people are likely to receive a diagnosis of depression, this study is likely to have missed cases of depression (particularly, mild cases of depression). However, we did search for people who were prescribed antidepressants (for the minimum adequate dosage period of six weeks) which should identify further cases of depression. A limitation of using prescribed drugs for identifying people with depression is that antidepressants could have been prescribed for other conditions such as sleep problems, particularly in older age where antidepressant prescribing is common.^25^ To reduce the likelihood of identifying people prescribed antidepressants for other conditions we searched for prescriptions of antidepressants that have been prescribed for six weeks or longer.

A further limitation is that adulthood environmental factors were only available for those patients who had a hospital admission, leading to missing data. This could bias our results be only capturing people with depression with other comorbidities in the models that adjusted for adulthood environmental factors. We also miss out on potentially important confounders, including education, and personal adulthood SES (e.g., occupation). As follow-up starts in 1980, we do not account for depression outcomes between childhood and mid-life. It is possible that individuals had experienced depression prior to follow-up (i.e., we can’t account for history of mental health conditions).

## Conclusions

In one of the largest studies of its kind, we report an association between higher risk for depression in older age and lower intelligence at age 11. This risk remained after adjusting for environmental factors experienced in childhood and adulthood.

## Supporting information

Record Checklist

## Data Availability

No data are available - data were accessed and analysed within eDRIS Safe Haven.

## Additional contributions

The authors would like to acknowledge the support of the eDRIS Team (Public Health Scotland) for their involvement in obtaining approvals, provisioning, and linking data and the use of the secure analytical platform within the National Safe Haven.

This work was part-funded by UKRI – Medical Research Council through the DATAMIND Hub (MRC reference: MR/W014386/1). The DATAMIND Hub and the authors would like to acknowledge the data providers who supplied the data sets enabling this research study.

## Author contributions

IJD, AMM, MHI, DMA, SRC designed the study and gained approvals for accessing these data. ELB, DMA, MHI contributed to data cleaning and analysis. ELB led on the analysis of this work. ELB produced the initial draft of this manuscript. All authors provided feedback on the statistical analysis and write up of this manuscript.

