## Supplementary for "Childhood intelligence and risk of depression in later-life: A longitudinal data-linkage study"

### Supplementary material

#### Supplement 1: Table of ICD codes for depression

| Code | Version | Code | Version |
|---|---|---|---|
| -29620 | icd9 | F320 | icd10 |
| -29621 | icd9 | F321 | icd10 |
| -29622 | icd9 | F322 | icd10 |
| -29623 | icd9 | F323 | icd10 |
| -29624 | icd9 | F324 | icd10 |
| -29625 | icd9 | F325 | icd10 |
| -29626 | icd9 | F328 | icd10 |
| -29630 | icd9 | F329 | icd10 |
| -29631 | icd9 | F330 | icd10 |
| -29632 | icd9 | F331 | icd10 |
| -29633 | icd9 | F332 | icd10 |
| -29634 | icd9 | F333 | icd10 |
| -29635 | icd9 | F334 | icd10 |
| -29636 | icd9 | F338 | icd10 |
| -29690 | icd9 | F339 | icd10 |
| -29699 | icd9 | F340 | icd10 |
| -3004 | icd9 | F341 | icd10 |
| -30112 | icd9 | F348 | icd10 |
| -3090 | icd9 | F349 | icd10 |
| -3091 | icd9 | F380 | icd10 |
| -3110 | icd9 | F381 | icd10 |
| -3111 | icd9 | F388 | icd10 |
| -3112 | icd9 | F39X | icd10 |
| -3113 | icd9 | F4320 | icd10 |
| -3114 | icd9 | F4321 | icd10 |
| -3115 | icd9 | | |
| -3116 | icd9 | | |
| -3117 | icd9 | | |
| -3118 | icd9 | | |
| -3119 | icd9 | | |
| -2962 | icd9 | | |
| -2963 | icd9 | | |
| -311 | icd9 | | |
| -309 | icd9 | | |

**Supplement 2: Table of antidepressants**

| Approved name |
|---|
| AGOMELATINE |
| AMITRIPTYLINE |
| AMITRIPTYLINE HYDROCHLORIDE WITH PERPHENAZINE |
| CITALOPRAM |
| CLOMIPRAMINE HYDROCHLORIDE |
| DOSULEPIN HYDROCHLORIDE |
| DOXEPIN |
| DULOXETINE |
| ESCITALOPRAM |
| FLUOXETINE |
| FLUPENTIXOL |
| FLUVOXAMINE MALEATE |
| IMIPRAMINE HYDROCHLORIDE |
| LOFEPRAMINE |
| MIANSERIN HYDROCHLORIDE |
| MIRTAZAPINE |
| MOCLOBEMIDE |
| NORTRIPTYLINE |
| PAROXETINE |
| PHENELZINE |
| REBOXETINE |
| SERTRALINE |
| TRAZODONE HYDROCHLORIDE |
| TRIMIPRAMINE |
| TRYPTOPHAN |
| VENLAFAXINE |
| VORTIOXETINE HYDROBROMIDE |

**Supplement 3: How many people had depression ICD codes reported in hospital admissions as the 'main' reason for admission and/or as an 'other' diagnosis?**

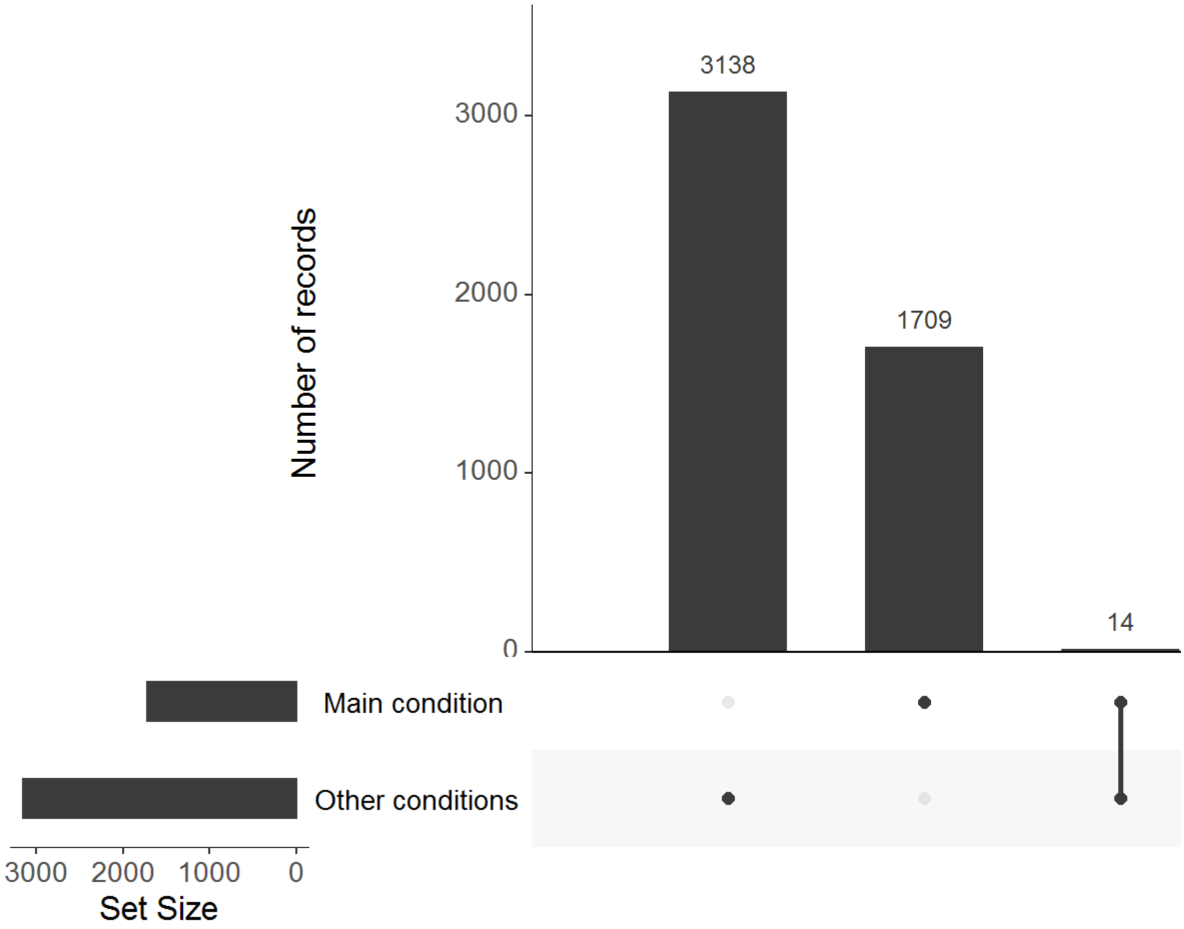


The plot shows the number of hospital admission records that had depression reported as the 'main' reason for a hospital admission, and/or depression reported as an 'other' condition when a participant was admitted to hospital. The dots show whether the diagnosis of depression was reported as a main and/or other condition. All hospital admission records that report diagnosis of depression are shown in the plot (i.e., one participant could have multiple hospital admissions).

### Supplement 4: Risk factors associated with depression after unadjusted and adjusted analyses

| Supplement 4: Risk factors associated with depression after unadjusted and adjusted analyses | | | | | | | | |
|---|---|---|---|---|---|---|---|---|
| | Unadjusted risk factors (Depression N/Total N) ± | | Model 1: Adjusted for childhood risk factors | | Model 2: Adjusted for adulthood risk factors | | Model 3: Adjusted for all risk factors | |
| | **CPH** HR (95% CI) | **Mixed effects model** | **CPH** HR (95% CI) | **Mixed effects model** | **CPH** HR (95% CI) | **Mixed effects model** | **CPH** HR (95% CI) | **Mixed effects model** |
| **Depression N/Total N (%)±** | N/A | N/A | 13022/49151 (26%) | 16595/96102 (17%) | 2776/11702 (24%) | 5637/57341 (10%) | 2757/11617 (24%) | 5537/56771 (10%) |
| **Male** | 0.65 (0.63 to 0.68) P<0.001 (n=14063/53037) | 0.68 (0.66 to 0.71) P<0.001 (n=18043/104762) | 0.64 (0.62 to 0.67) P<0.001 | 0.68 (0.65 to 0.71) P<0.001 | 0.71 (0.66 to 0.77) P<0.001 | 0.84 (0.77 to 0.92) P<0.001 | 0.71 (0.66 to 0.77) P<0.001 | 0.84 (0.77 to 0.92) P<0.001 |
| **Moray House Test** | 0.89 (0.87 to 0.90) P<0.001 (n=13118/49506) | 0.95 (0.93 to 0.97) P<0.001 (n=16781/96960) | 0.88 (0.86 to 0.89) P<0.001 | 0.94 (0.92 to 0.96) P<0.001 | 0.91 (0.88 to 0.95) P<0.001 | 0.95 (0.91 to 1.00) P=0.035 | 0.92 (0.88 to 0.95) P<0.001 | 0.95 (0.91 to 1.00) P=0.032 |
| **Position in family** | 1.03 (1.02 to 1.03) P<0.001 (n=13934/52542) | 1.01 (1.00 to 1.02) P=0.142 (n=17813/103605) | N/A | N/A | N/A | N/A | N/A | N/A |
| **Size of family** | 1.07 (1.05 to 1.09) P<0.001 (n=13931/52538) | 1.03 (1.01 to 1.05) P=0.012 (n=17810/103591) | 1.02 (1.00 to 1.04) P=0.013 | 1.00 (0.98 to 1.03) P=0.709 | N/A | N/A | 1.02 (0.98 to 1.06) P=0.338 | 1.00 (0.96 to 1.05) P=1.000 |
| **Carstairs** | 1.03 (1.01 to 1.04) P<0.001 (n=3047/12760) | 1.01 (1.00 to 1.03) P=0.048 (6165/63163) | N/A | N/A | 1.02 (1.01 to 1.03) P=0.006 | 1.01 (1.00 to 1.03) P=0.200 | 1.02 (1.00 to 1.03) P=0.008 | 1.01 (0.99 to 1.03) P=0.252 |
| **Remote location*** | 0.92 (0.83 to 1.01) P=0.088 (n=3022/12650) | 1.05 (0.94 to 1.18) P=0.350 (n=6117/62376) | N/A | N/A | 0.98 (0.88 to 1.09) P=0.722 | 1.09 (0.97 to 1.22) P=0.165 | 0.98 (0.89 to 1.09) P=0.769 | 1.09 (0.97 to 1.22) P=0.160 |
| **SIMD – Linear** | 1.32 (1.18 to 1.49) P<0.001 (n=3029/12692) | 1.12 (0.99 to 1.28) P=0.077 (n=6127/62649) | N/A | N/A | N/A | N/A | N/A | N/A |

CI: confidence interval; CPH: cox proportional hazards; HR: hazards ratio; SIMD: Scottish Index of Multiple Deprivation
*Accessible Rural Areas, Remote Rural Areas, Very Remote Rural Areas
± CPH analyses is based on number of people in analysis, whereas, mixed effects models are based on number of observations
Cox proportional hazards: one record for each participant. The earliest diagnosis of depression. When a person had a hospital admission(s) for depression and were prescribed antidepressants, we included the earliest diagnosis of depression that had adult environmental factors reported.
The number of records included in each statistical model varies because records with missing environmental factors are excluded from the analyses.

### Supplement 5: Sex stratified analyses

| Supplement 5: Risk factors associated with depression after sex-stratified analyses | | | | | | | | |
|---|---|---|---|---|---|---|---|---|
| | **Sex-stratified risk factors (Depression N/Total N) ±** | | **Model 1: Adjusted for childhood risk factors** | | **Model 2: Adjusted for adulthood risk factors** | | **Model 3: Adjusted for all risk factors** | |
| | **CPH** HR (95% CI) | **Mixed effects model** | **CPH** HR (95% CI) | **Mixed effects model** | **CPH** HR (95% CI) | **Mixed effects model** | **CPH** HR (95% CI) | **Mixed effects model** |
| **Depression N/Total N (%)±** | N/A | N/A | 13022/49151 (26%) | 16595/96102 (17%) | 2776/11702 (24%) | 5637/57341 (10%) | 2757/11617 (24%) | 5537/56771 (10%) |
| **Moray House Test** | 0.87 (0.86 to 0.89) P<0.001 (n=13118/49506) | 0.94 (0.92 to 0.96) P<0.001 (n=16781/96960) | 0.88 (0.86 to 0.89) P<0.001 | 0.94 (0.92 to 0.96) P<0.001 | 0.91 (0.88 to 0.95) P<0.001 | 0.95 (0.91 to 1.00) P=0.035 | 0.92 (0.88 to 0.96) P<0.001 | 0.95 (0.91 to 1.00) P=0.032 |
| **Position in family** | 1.02 (1.01 to 1.03) P<0.001 (n=13934/52542) | 1.01 (1.00 to 1.02) P=0.271 (n=17813/103605) | N/A | N/A | N/A | N/A | N/A | N/A |
| **Size of family** | 1.06 (1.05 to 1.08) P<0.001 (n=13931/52538) | 1.02 (1.00 to 1.04) P=0.026 (n=17810/103591) | 1.02 (1.00 to 1.04) P=0.013 | 1.00 (0.98 to 1.03) P=0.709 | N/A | N/A | 1.02 (0.98 to 1.06) P=0.334 | 1.00 (0.96 to 1.05) P=1.000 |
| **Carstairs** | 1.03 (1.01 to 1.04) P<0.001 (n=3047/12760) | 1.01 (1.00 to 1.03) P=0.044 (n=6165/63163) | N/A | N/A | 1.02 (1.01 to 1.03) P=0.006 | 1.01 (1.00 to 1.03) P=0.193 | 1.02 (1.00 to 1.03) P=0.008 | 1.01 (0.99 to 1.03) P=0.246 |
| **Remote location*** | 0.92 (0.84 to 1.02) P=0.118 (n=3022/12650) | 1.06 (0.95 to 1.18) P=0.289 (n=6117/62376) | N/A | N/A | 0.98 (0.88 to 1.09) P=0.712 | 1.09 (0.97 to 1.22) P=0.154 | 0.98 (0.88 to 1.09) P=0.760 | 1.09 (0.97 to 1.22) P=0.145 |
| **SIMD – Linear** | 1.35 (1.20 to 1.52) P<0.001 (n=3029/12692) | 1.13 (1.00 to 1.28) P=0.058 (n=6127/62649) | N/A | N/A | N/A | N/A | N/A | N/A |

CI: confidence interval; CPH: cox proportional hazards; HR: hazards ratio
± CPH analyses is based on number of people in analysis, whereas, mixed effects models are based on number of observations
*Accessible Rural Areas, Remote Rural Areas, Very Remote Rural Areas

### Supplement 6: Unadjusted mixed effect models performed on separate populations of females and males

| Supplement 6: Unadjusted mixed effect models performed on separate populations of females and males | | |
|---|---|---|
| | **Females** | **Males** |
| | **Mixed effects model (Depression N observations/Total N observations)** | **Mixed effects model (Depression N observations/Total N observations)** |
| | HR (95% CI) | HR (95% CI) |
| **Moray House Test** | 0.94 (0.92 to 0.97) P<0.001 (n=10618/50383) | 0.93 (0.90 to 0.96) P<0.001 (n=6163/46577) |
| **Position in family** | 1.00 (0.99 to 1.02) P=0.646 (n=11303/53589) | 1.01 (0.99 to 1.03) P=0.228 (n=6510/50016) |
| **Size of family** | 1.02 (1.00 to 1.05) P=0.116 (n=11301/53582) | 1.03 (0.99 to 1.06) P=0.113 (n=6509/50009) |
| **Carstairs** | 1.02 (1.00 to 1.04) P=0.030 (n=3859/32898) | 1.00 (0.98 to 1.02) P=0.779 (n=2306/30265) |
| **Remote location*** | 1.05 (0.90 to 1.22) P=0.551 (n=3829/32432) | 1.08 (0.92 to 1.27) P=0.353 (n=2288/29944) |
| **SIMD – Linear** | 1.23 (1.04 to 1.46) P=0.018 (n=3836/32572) | 0.98 (0.80 to 1.20) P=0.839 (n=2291/30077) |
| CI: confidence interval; HR: hazards ratio; SIMD: Scottish Index of Multiple Deprivation<br>*Accessible Rural Areas, Remote Rural Areas, Very Remote Rural Areas | | |

### Supplement 7: Risk factors associated with depression after unadjusted and adjusted analyses. Outcome is depression identified in prescribed drugs records only (not hospital admissions data)

| Supplement 7: Risk factors associated with depression after unadjusted and adjusted analyses. Outcome is depression identified in prescribed drugs records only (not hospital admissions data) | | | | |
|---|---|---|---|---|
| | **Unadjusted risk factors** | **Model 1: Adjusted for childhood risk factors** | **Model 2: Adjusted for adulthood risk factors** | **Model 3: Adjusted for all risk factors** |
| | **Mixed effects model (Depression N observations/Total N observations)** | **Mixed effects model** | **Mixed effects model** | **Mixed effects model** |
| **Depression N/Total N (%)** | N/A | 12214/96102 (13%) | 1225/57341 (2%) | 1219/56771 (2%) |
| **Male** | 0.72 (0.70 to 0.75) P<0.001 (n=13182/104762) | 0.72 (0.70 to 0.72) P<0.001 | 1.05 (0.94 to 1.18) P=0.410 | 1.04 (0.93 to 1.07) P=0.527 |
| **Moray House Test** | 1.01 (1.00 to 1.03) P=0.145 (n=12306/96960) | 1.00 (0.98 to 1.02) P=0.100 | 1.01 (0.95 to 1.07) P=0.760 | 1.01 (0.95 to 1.07) P=0.844 |
| **Position in family** | 0.99 (0.98 to 1.00) P=0.171 (n=13058/103605) | N/A | N/A | N/A |
| **Size of family** | 0.99 (0.97 to 1.01) P=0.326 (n=13055/103591) | 0.99 (0.97 to 1.01) P=0.268 | N/A | 0.99 (0.94 to 1.05) P=0.813 |
| **Carstairs** | 0.97 (0.95 to 0.99) P<0.001 (n=1345/63163) | N/A | 0.97 (0.95 to 0.99) P=0.001 | 0.97 (0.95 to 0.99) P=0.002 |
| **Remote location*** | 1.16 (1.00 to 1.35) P=0.046 (n=1326/62376) | N/A | 1.12 (0.96 to 1.31) P=0.151 | 1.13 (0.96 to 1.32) P=0.140 |
| **SIMD – Linear** | 1.12 (0.99 to 1.28) P=0.077 (n=1332/62649) | N/A | N/A | N/A |
| CI: confidence interval; HR: hazards ratio; SIMD: Scottish Index of Multiple Deprivation<br>*Accessible Rural Areas, Remote Rural Areas, Very Remote Rural Areas | | | | |

### Supplement 8: Risk factors associated with depression after unadjusted and adjusted analyses. Outcome is depression identified in hospital admissions data only (not prescribed drugs data)

| Supplement 8: Risk factors associated with depression after unadjusted and adjusted analyses. Outcome is depression identified in hospital admissions data only (not prescribed drugs data) | | | | |
|---|---|---|---|---|
| | **Unadjusted risk factors** | **Model 1: Adjusted for childhood risk factors** | **Model 2: Adjusted for adulthood risk factors** | **Model 3: Adjusted for all risk factors** |
| | **Mixed effects model (Depression N observations/Total N observations)** | **Mixed effects model** | **Mixed effects model** | **Mixed effects model** |
| **Depression N/Total N (%)** | N/A | 4381/96102 (4.6%) | 4412/57341 (7.7%) | 4318/56771 (7.6%) |
| **Male** | 0.78 (0.71 to 0.86) P<0.001 (n=4861/104762) | 0.78 (0.71 to 0.86) P<0.001 | 0.79 (0.72 to 0.88) P<0.001 | 0.79 (0.71 to 0.88) P<0.001 |
| **Moray House Test** | 0.86 (0.82 to 0.90) P<0.001 (n=4475/96960) | 0.86 (0.81 to 0.90) P<0.001 | 0.95 (0.90 to 1.00) P=0.031 | 0.94 (0.89 to 0.99) P=0.026 |
| **Position in family** | 1.03 (1.00 to 1.05) P=0.045 (n=4755/103605) | N/A | N/A | N/A |
| **Size of family** | 1.08 (1.03 to 1.13) P=0.002 (n=4755/103591) | 1.03 (0.98 to 1.08) P=0.265 | N/A | 1.00 (0.95 to 1.05) P=0.910 |
| **Carstairs** | 1.02 (1.01 to 1.04) P=0.003 (n=4820/63163) | N/A | 1.02 (1.00 to 1.04) P=0.022 | 1.02 (1.00 to 1.04) P=0.031 |
| **Remote location*** | 1.04 (0.92 to 1.18) P=0.535 (n=4791/62376) | N/A | 1.08 (0.94 to 1.23) P=0.272 | 1.08 (0.95 to 1.23) P=0.259 |
| **SIMD – Linear** | 1.12 (0.99 to 1.28) P=0.077 (n=4795/62649) | N/A | N/A | N/A |
| CI: confidence interval; HR: hazards ratio; SIMD: Scottish Index of Multiple Deprivation<br>*Accessible Rural Areas, Remote Rural Areas, Very Remote Rural Areas | | | | |
